# Assessing responsiveness to direct verbal suggestions in depersonalization-derealization disorder

**DOI:** 10.1101/2022.01.21.22269634

**Authors:** L. S. Merritt Millman, Elaine C. M. Hunter, Anthony S. David, Guido Orgs, Devin B. Terhune

## Abstract

The dissociative disorders and germane conditions are reliably characterized by elevated responsiveness to direct verbal suggestions. However, it remains unclear whether atypical responsiveness to suggestion is similarly present in depersonalization-derealization disorder (DDD). 55 DDD patients and 36 healthy controls completed a standardised behavioural measure of direct verbal suggestibility that includes a correction for compliant responding (BSS-C), and psychometric measures of depersonalization-derealization (CDS), mindfulness (FFMQ), imagery vividness (VVIQ), and anxiety (GAD-7). Patients displayed nonsignificantly lower suggestibility than controls, (*g* = 0.26) but significantly lower mindfulness (*g* = 1.38), and imagery vividness (*g* = 0.63), and significantly greater anxiety (*g* = 1.39). Although suggestibility did not correlate with severity of depersonalization-derealization symptoms in controls, *r*=-.03 [95% CI: -.36, .30], there was a weak tendency for a positive association in patients, *r*=.25, [95% CI: -.03, .48]. Exploratory analyses revealed that patients with more severe anomalous bodily experiences were also more responsive to suggestion, an effect not seen in controls. This study demonstrates that DDD is not characterized by elevated responsiveness to direct verbal suggestions. These results have implications for the aetiology and treatment of this condition, as well as its classification as a dissociative disorder in psychiatric nosology.

## 1 Introduction

*Dissociation*, broadly defined, may manifest as a disconnection from, or alteration of, one’s identity, consciousness and memory (DSM-5^1^), that is typically characterized by an attenuation in, or disruption of, the integration of mental processes.^2^ It has become increasingly evident that, within this broad constellation of symptoms, most dissociative experiences can be considered to reflect either *compartmentalization* or *detachment*.^3^ These categories encompass different symptoms and clinical conditions, and are hypothesised to arise from independent mechanisms.^3-5^ Compartmentalization symptoms involve the fragmentation of processes that are normally integrated, such as dissociative amnesia, identity disturbances and functional neurological symptoms.^3,4,6^ By contrast, detachment symptoms are characterized by disruptions in the integration of conscious awareness including discontinuities in experience and the perceived separation from the self, body, and one’s surroundings.^3,4^

Within the Diagnostic and Statistical Manual of Mental Disorders (DSM-5^1^), *dissociative amnesia, dissociative identity disorder*, and *depersonalization-derealization disorder* (DDD) are classified within the category of dissociative disorders. Whilst the former two are typified by compartmentalization symptoms,^3,4^ DDD is primarily characterized by detachment pertaining to the self (depersonalization) or to one’s environment (derealization). This symptom demarcation amounts to a fissure within dissociative psychopathology and places DDD in a unique position with regard to other dissociative disorders. Indeed, a recent meta-analysis^2^ demonstrated that patients with DDD score lower on the *Dissociative Experiences Scale* (DES^7^) than those diagnosed with dissociative identity disorder and other dissociative disorders as well as those with posttraumatic stress disorder (PTSD), borderline personality disorder, and functional neurological disorder, which are not classified as dissociative disorders.^1^ The symptom differences seen in DDD as compared to other dissociative disorders are potentially attributable to differential aetiologies: whereas trauma exposure is considered the sole antecedent of dissociative disorders and PTSD,^8-11^ it seems to only be implicated in a minority of DDD cases.^12-14^ These differential patterns are further corroborated by a recent systematic review, which reported that patients with DID and the dissociative subtype of PTSD displayed greater neurophysiological similarity than either group displayed with DDD patients.^15^ Collectively, these disparate lines of research strongly suggest that DDD is distinct from the dissociative disorders, with differing phenomenology, aetiology, and mechanisms.

The capacity to respond to direct verbal suggestions (suggestibility) provides a potential route to further elucidate how DDD fits within the dissociative disorders taxonomy. Hypnotic suggestibility, which is characterized by pronounced distortions in the sense of agency,^16,17^ and dissociation are historically intertwined^18-20^ and have long been theorized to have overlapping mechanisms.^21-23^ A recent meta-analysis^24^ found moderate-to-large effect sizes of elevated hypnotic suggestibility relative to controls in dissociative identity disorder and mixed dissociative disorders, and two germane conditions (trauma and stressor-related disorders and functional neurological disorder).^25-27^ Moreover, the available evidence suggests that elevated suggestibility is selective to dissociative psychopathology as it is not observed in anxiety disorders^29^ or schizophrenia.^30,31^ If it is a cognitive feature of generalized dissociative psychopathology, DDD would be expected to be associated with elevated suggestibility. In addition, we would expect that depersonalization-derealization symptom severity would positively scale with suggestibility, as observed in other conditions.^32^ By contrast, responsiveness to verbal suggestions is often conceptualized as a form of compartmentalization wherein one’s actions and perceptual states are separated from the antecedent intentions that produced the corresponding responses.^3,4^ A corollary of this view is that suggestibility will selectively accompany compartmentalization symptomatology and thus should not be observed in DDD.^24,33^ The factors that moderate this association remain unclear, with mindfulness and imagery as two potential candidates. Previous research suggests reduced mindfulness or metacognition in highly suggestible individuals^34-38^ and in dissociative disorders,^36,39,40^ as well as an impaired ability to generate visual images in DDD.^41^ This research points towards the importance of examining both of these factors in the context of suggestibility in DDD.

This study sought to discriminate between the competing predictions that DDD patients would display greater suggestibility than controls or that the two groups would display comparable levels of suggestibility. DDD patients and non-clinical controls completed a standardized behavioural measure of direct verbal suggestibility and psychometric measures of depersonalization-derealization, mindfulness, and imagery vividness. We further evaluated whether depersonalization-derealization symptomatology would moderate any group difference, with the expectation that symptom severity would be associated with greater suggestibility. We also expected that mindfulness would moderate the group differences, with greater suggestibility associated with poorer mindfulness, particularly in the DDD group. Finally, exploratory analyses were conducted to examine whether imagery and anxiety may also play a role in the group differences.

## 2 Methods

### 2.1 Studies

Patients and controls were drawn from in-person and online variants of a larger study measuring bodily awareness in DDD. The in-person variant was interrupted in March 2020 due to the COVID-19 pandemic, leading to the implementation of the online variant.

### 2.2 Samples

Participants with DDD were recruited through the Depersonalization Research Unit at King’s College London from among those who had previously expressed a willingness to participate in research; a post advertising these studies (in-person and online) on the UK DDD charity website (https://www.unrealuk.org/); social media channels; relevant email lists; and thedepersonalisationclinic.com. Healthy, age-matched controls were recruited through advertisements, newsletters, and social media. Interested participants were given a detailed information sheet before taking part in a phone screening to assess eligibility. All eligible participants provided informed consent in accordance with the Declaration of Helsinki and Goldsmiths, University of London ethical approval and were compensated £40 for completion of both phases of the larger study (see pre-registrations on OSF: https://osf.io/4brch/?view_only=f429afb10a52489aac7e5110663539a8, https://osf.io/efz53/?view_only=e90a5e3fa025429fa2a5637bf30c6102).

Patients and controls were included if they met the following criteria: aged 18-70; no previous or current head injury; no severe drug or alcohol use; no neurological disorder; and no severe physical impairment affecting motor performance. DDD patients were required to meet DSM-5 diagnostic criteria^1^ for current DDD including: having persistent (either chronic or recurrent) episodes of depersonalization and derealization; being aware that their symptoms are a subjective experience; the symptoms cause distress and/or impairment to their functioning; and the symptoms are not better explained by another disorder or substance use. In addition, DDD patients were also required to have no self-reported comorbid current diagnosis of schizophrenia, other psychotic disorder, or PTSD. Control participants were required to not meet DSM-5 criteria for DDD and have no other self-reported current psychiatric diagnoses. These criteria were assessed as part of a structured telephone screening interview. To take part in the online study, participants could be residing anywhere worldwide whereas to take part in the in-person study, participants were required to be currently living in London or with access to the city of London.

This study was part of two larger studies on bodily awareness in DDD, which each included planned sample sizes of 30 DDD patients and 30 controls on the basis of an *a priori* power analysis (see pre-registrations on OSF: https://osf.io/4brch/?view_only=f429afb10a52489aac7e5110663539a8, https://osf.io/efz53/?view_only=e90a5e3fa025429fa2a5637bf30c6102). 57 patients and 39 controls consented to participate, but 2 patients and 3 controls dropped out post-baseline completion and therefore their data was excluded from these analyses. The final sample for the present study included 55 DDD patients and 36 controls, which allowed us to detect group differences corresponding to Cohen’s *d* ≥ .61 (two-tailed, α =.05, power=.80; conducted using G*Power 3.1^42^) based on a t-test sensitivity analysis.

### 2.3 Measures

The *Cambridge Depersonalization Scale* (CDS^43^) is a 29-item self-report measure of depersonalization and derealization experiences. Respondents rate the frequency (0 [“never”] – 4 [“all the time”]) and duration (1 [“few seconds”] – 6 [“more than a week”]) of different experiences in the preceding six months. If 0 (“never”) is endorsed for frequency, a score of 0 is also inferred for duration. As the original study from which these self-reports are drawn concerned week-to-week changes in symptoms, respondents completed the measure with reference to their experiences in the preceding week. Frequency and duration scores are summed with a total scoring range of 0-290 (Cronbach’s α = 0.96). The cut-off score for a clinical diagnosis of DDD in 80% of cases is 70.^43^ Scores were also calculated for four subscales: emotional numbing (CDS-EN, 6 items; α = 0.86), anomalous body experience (CDS-ABE, 9 items; α = 0.91), anomalous subjective recall (CDS-ASR, 5 items; α = 0.82), and alienation from surroundings (CDS-AfS, 4 items; α = 0.91).^44^

The *Brief Suggestibility Scale* (BSS^11^) is a computerized behavioural scale used to measure non-hypnotic direct verbal suggestibility. This scale has been shown to moderately correlate with a standardized measure of hypnotic suggestibility.^11^ Respondents are aurally presented with six verbal suggestions for arm heaviness, a dream, hands moving together, an inability to open eyes, arm rigidity, and a music hallucination followed by simple behavioural tests. Respondents subsequently rate the extent to which they had responded to each suggestion according to suggestion-specific behavioural descriptions using a continuous visual analogue scale from 0 (no response) to 1 (complete response) followed by a 6-point Likert-scale rating of perceived involuntariness of each response (0 = “did not experience at all”; 1 [voluntary] to 5 [involuntary]),^45^ in order to capture the classic suggestion effect^46^ and correct for compliant responses.^47^ Both the behavioural and involuntariness measures (6-item means) displayed good internal consistency (αs = 0.72, 0.72, respectively). Scores were corrected for compliance by computing the mean of z-transformed behavioural and involuntariness scores (BSS-C^11^).

The *Five Facet Mindfulness Questionnaire* (FFMQ^48^) is a 39-item scale measuring five dimensions of mindfulness: Observing, Describing, Acting with Awareness, Non-Judging, and Non-Reactivity. Items are rated on a Likert scale of 1 (“never or very rarely true”) to 5 (“very often or always true”). As is the case with the CDS, respondents completed this scale with reference to the preceding week. Total scores range from 39-195, with higher scores reflecting increased mindfulness, and subscale scores ranging from 8-40, or 7-35 (Non-reactivity). We were primarily interested in the Acting with Awareness subscale because of the phenomenological similarity with involuntary responses to suggestions; a representative item includes “It seems I am ‘running on automatic’ without much awareness of what I’m doing” (reverse-scored). The FFMQ displayed high internal consistency overall (α = 0.92) and for each subscale: Observing (FFMQ-O, 8 items; α = 0.81), Describing (FFMQ-D, 8 items; α = 0.87), Acting with Awareness (FFMQ-AA, 8 items; α = 0.92), Non-Judging (FFMQ-NJ, 8 items; α = 0.94), and Non-Reactivity (FFMQ-NR, 7 items; α = 0.82).

The *Vividness of Visual Imagery Questionnaire* (VVIQ^49^) is a 16-item scale measuring the intensity of imagined visual scenes. The items comprise four groups involving a specific scenario (e.g., “Think of the front of a shop which you often go to. Consider the picture that comes before your mind’s eye”), in response to which participants rate the vividness of specific details within each scenario using a five-point Likert scale (1: “perfectly clear and vivid as normal vision” to 5: “no image at all, you only “know” that you are thinking of the object”) with scores ranging from 16-80. This scale displayed high internal consistency (α = 0.94).

The *Generalized Anxiety Disorder - 7* (GAD-7^50^) is a brief self-report scale of generalized anxiety. The 7 items ask about symptoms over the last two weeks and are rated from 0 (“not at all”) to 3 (“nearly every day”) with total scores ranging from 0-21. The cut-off points for mild, moderate, and severe anxiety are 5, 10, and 15, respectively.^51^ A score of 10 or greater acts as the single screening cut-off point with a sensitivity of 89% and a specificity of 82% for GAD.^50^ This scale displayed strong internal consistency (α = .91).

### 2.4 Procedure

After a telephone interview and screening to ensure eligibility, and providing informed consent, the BSS, VVIQ and GAD-7 were administered to all participants online via Qualtrics (www.qualtrics.com) as part of a battery of measures. Participants in the online study were then sent the CDS and FFMQ via Qualtrics and asked to complete them prior to their first online behavioural session of the larger study whereas participants in the in-person study completed the CDS and FFMQ during their first in-person session of the larger study. A debrief was provided to all participants after completion of the study.

### 2.5 Statistical Analyses

All data were analysed using *R* (Version 4.1.0).^52^ There were no missing data for the in-person participants and in the case of missing data at Time 1 for the online participants, expectation maximisation was used to estimate any missing data as part of the larger study. There were no missing data for the VVIQ, BSS-C or GAD-7 at baseline, or for the FFMQ at Time 1, and missing data for the CDS at Time 1 was found for 1.5%-5.9% of cases. Little’s MCAR test was non-significant, χ^2^ (552) = .00, *p* = 1.00, and therefore we assume the data were missing completely at random. The data were normally distributed, as evaluated with QQ plots and Shapiro-Wilk tests, with assumptions of homogeneity of variance met on all measures except for the CDS. One patient was identified as an outlier (*M* ± 2.5 *SD*s) on the CDS; their score was winsorized to allow for inclusion in the final analyses. The two groups were compared on demographics and psychometric measures using between-groups Welch ANOVAs (DDD vs. controls), with Hedges’ *g* as a measure of effect size, and Chi-squared tests. A complementary Bayesian t-test (BF_10_, default Cauchy prior = .707) was also conducted with BSS-C scores. Next, we performed two moderation analyses on BSS-C scores with Group as a predictor and, alternately, CDS scores and FFMQ-AA subscale scores as moderators. Pearson correlations were computed to assess associations between mindfulness (FFMQ) and suggestibility (BSS-C) in each group separately and the collapsed total sample. Exploratory analyses investigated associations between CDS and FFMQ subscales, VVIQ, GAD-7 and BSS-C scores. All analyses were two-tailed (α < .05) except the exploratory correlational analyses which used a lower threshold for significance (α < .01).

## 3 Results

### 3.1 Patient and control demographics

As can be seen in Table 1, patients with DDD and controls were relatively well matched on the demographic variables, with a weak trend toward lower education in the former group. Two DDD (4%) patients scored below the recommended clinical cutoff of 70 on the CDS,^43^ with the remainder of patients scoring above this threshold. By contrast, only two participants in the control group (6%) scored above this threshold. In turn, patients with DDD and controls significantly differed on CDS scores (**Table 1** and **Figure 1**).

**Table 1.** Demographic characteristics and research variables in patients with DDD and controls.

| Variable | DDD (n=55) | Control (n=36) | $\chi^2$ | *p* | $\Phi$ |
| --- | --- | --- | --- | --- | --- |
|  | % (n) | % (n) |  |  |  |
| Education (% university) | 62<br>(34) | 81<br>(29) | 2.76 | .096 | .17 |
| Employment (% employed) | 56<br>(31) | 47<br>(17) | 0.41 | .52 | .07 |
| Gender (% female) | 65<br>(36) | 75<br>(27) | 0.54 | .46 | .08 |
| Location (% in UK) | 76<br>(42) | 78<br>(28) | 0.00 | 1.00 | .00 |
|  | *M (SD)* | *M (SD)* | *F (df)* | *p* | *g* |
| Age | 34.9<br>(13.2) | 32.5<br>(11.3) | 0.87<br>(1, 82.5) | .36 | 0.19 |
| CDS | 149<br>(43.3) | 30.2<br>(20.3) | 309.00<br>(1, 82.1) | <.001 | 3.27 |
| BSS-C | -0.19<br>(1.8) | 0.29<br>(2.0) | 1.38<br>(1, 69.6) | .24 | 0.26 |
| FFMQ | 105<br>(19.3) | 131<br>(17.6) | 42.20<br>(1, 79.8) | <.001 | 1.38 |
| VVIQ | 43.8<br>(14.6) | 52.4<br>(11.2) | 9.86<br>(1, 86.7) | .002 | 0.63 |
| GAD-7 | 10.7<br>(5.60) | 4.03<br>(3.05) | 54.3<br>(1, 86.6) | <.001 | 1.39 |

*Notes.* DDD = depersonalization-derealization disorder; CDS = *Cambridge Depersonalization Scale*; BSS-C = *Brief Suggestibility Scale-Composite*; FFMQ = *Five Facet Mindfulness Questionnaire*; VVIQ = *Vividness of Visual Imagery Questionnaire*.

**Figure 1.**
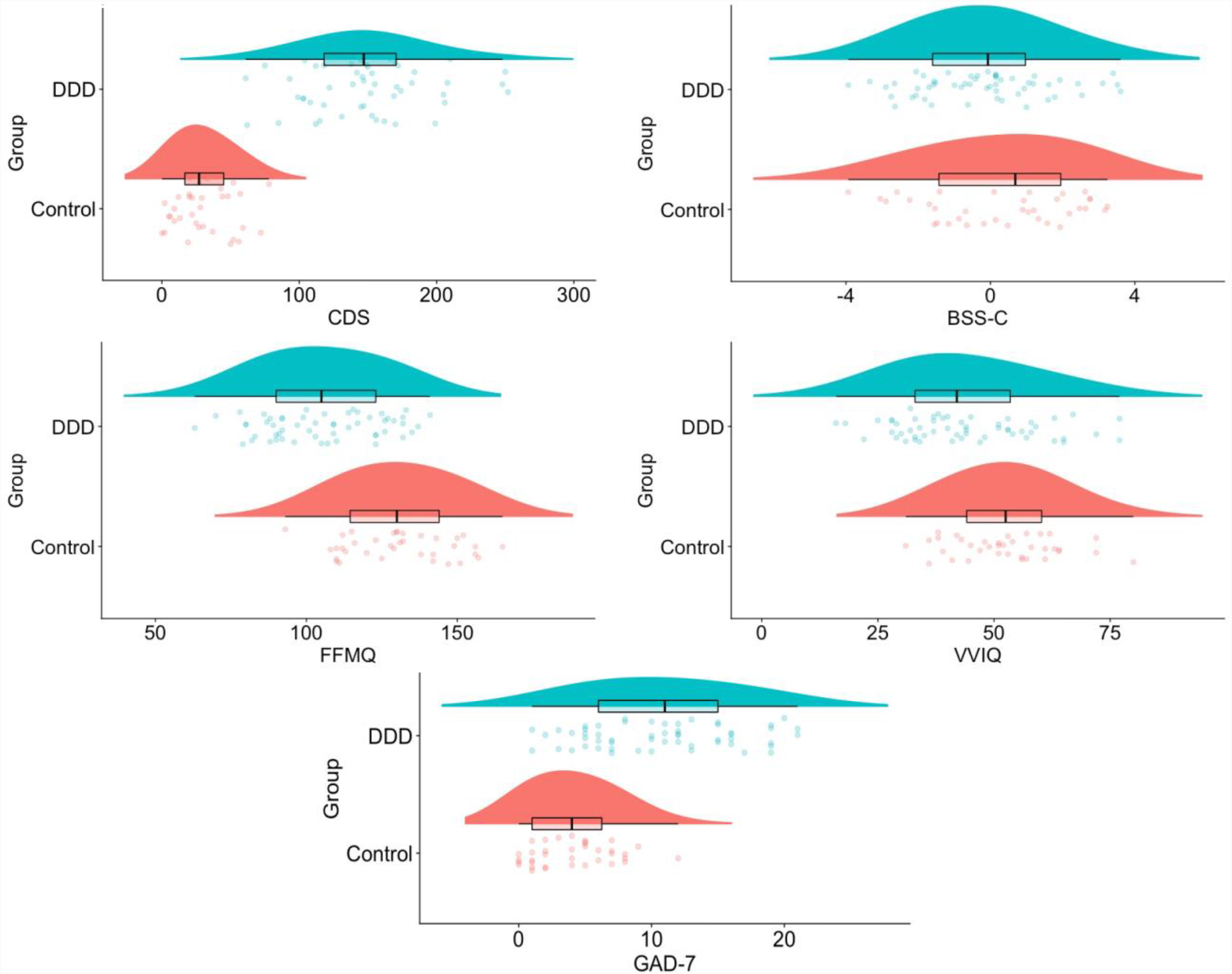
Research variables as a function of group (DDD: *n* = 55; Control: *n* = 36) *Notes*. DDD = depersonalization-derealization disorder; CDS = *Cambridge Depersonalization Scale*; BSS-C = *Brief Suggestibility Scale-Composite*; FFMQ = *Five Facet Mindfulness Questionnaire*; VVIQ = *Vividness of Visual Imagery Questionnaire*; GAD-7 = *Generalized Anxiety Disorder – 7*.

**Figure 2.**
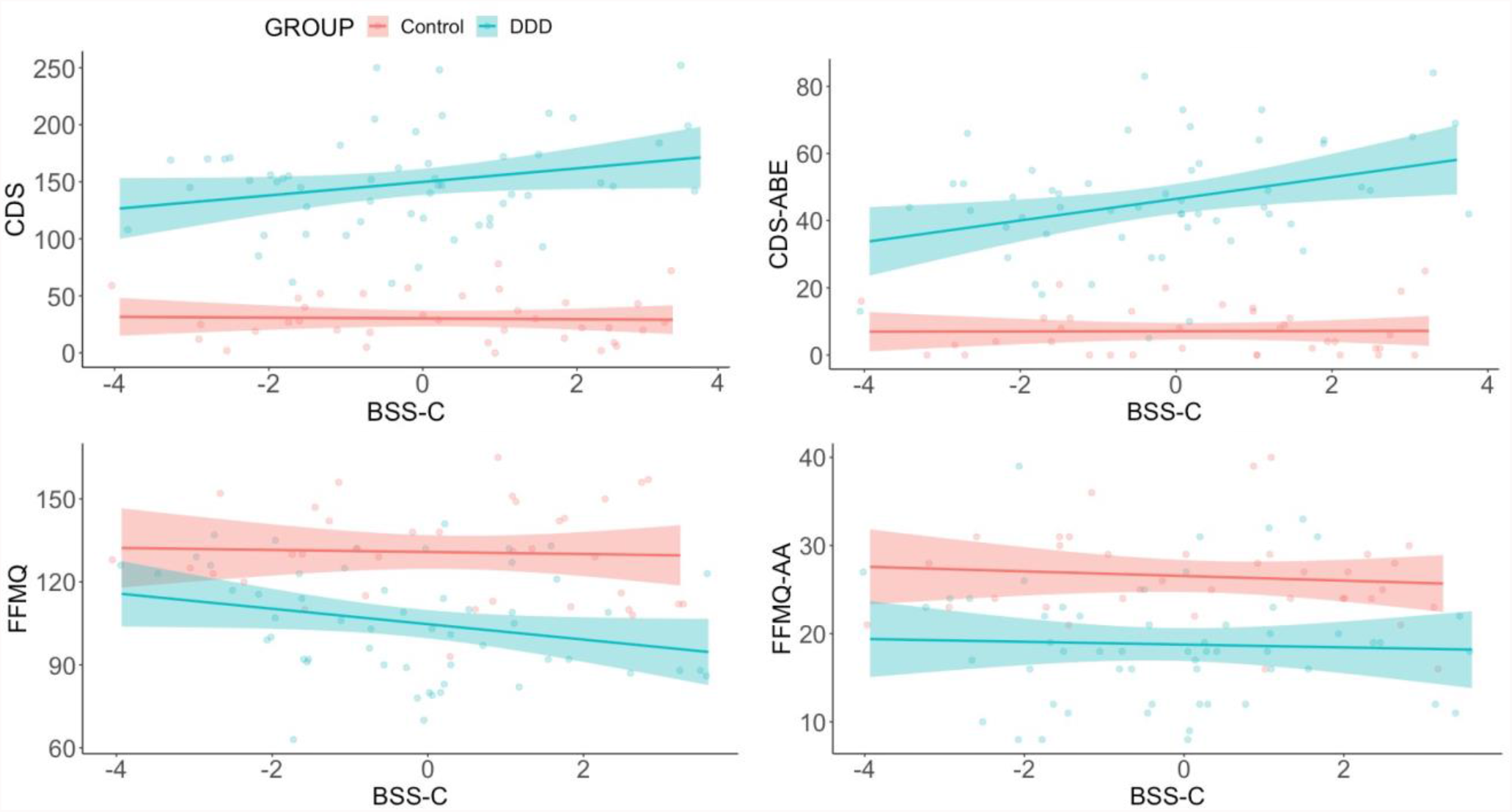
Correlations between suggestibility, depersonalization, and mindfulness (DDD: *n* = 55; control: *n* = 36) *Notes*. DDD = depersonalization-derealization disorder; CDS = *Cambridge Depersonalization Scale*; BSS-C = *Brief Suggestibility Scale-Composite*; FFMQ = *Five Facet Mindfulness Questionnaire*; CDS-ABE = *Cambridge Depersonalization Scale – Anomalous Bodily Experience*; FFMQ-AA = *Five Facet Mindfulness Questionnaire – Acting with Awareness*

### 3.2 Responsiveness to suggestions

As can be seen in **Table 1** and **Figure 1**, patients with DDD displayed non-significantly lower suggestibility (BSS-C) than controls, with a small effect size (*g* = 0.26). A complementary Bayesian *t*-test using a default prior yielded moderate evidence in favour of the null hypothesis, BF_10_ = .11. This suggests that DDD patients and healthy controls were relatively comparable in direct verbal suggestibility, but the results are insensitive with regard to whether patients were lower in suggestibility than controls. This result is at odds with the prediction that DDD patients would be more responsive to direct verbal suggestions.

### 3.3 Responsiveness to suggestion and CDS severity

One interpretation of the lack of a robust difference in suggestibility between patients and controls is that such a Group effect is moderated by depersonalization-derealization symptomatology, that is, atypical suggestibility is specific to patients with a more severe symptom profile. We evaluated this possibility by assessing whether CDS scores would moderate the association between Group and suggestibility (BSS-C). The overall model was non-significant, *F*(3, 87) = 1.52, *p* = .21, with non-significant Group x CDS interaction, *b* = .01, *t*(87) = 0.82, *p* = .42, and CDS effects, *b* = -.00, *t*(87) = -.21, *p* = .83, although there was a weak trend toward a Group effect, *b* = -2.11, *t*(87) = -1.97, *p* = .051, with patients with DDD displaying marginally lower BSS-C scores. Although this analysis suggests that the association between depersonalization-derealization symptoms and suggestibility did not differ between groups, Pearson correlation analyses revealed a suggestive effect in patients (see **Figure 3**). In the total collapsed sample, the association between CDS and BSS-C scores was near-zero, *r*(89) = -.02, *p* = .83 [95% CI: -.23, .18], and this held in the controls, *r*(34) = -.03, *p* = .84 [95% CI: -.36, .30]. By contrast, in the patients, there was a weak trend towards a positive correlation, *r*(53) = .25, *p* = .07 [95% CI: -.03, .48], though these two group correlations did not significantly differ, *z* = 1.28, *p* = .20. Taken together, these results provide preliminary evidence that responsiveness to verbal suggestions scales with symptom severity in patients with DDD.

**Figure 3.**
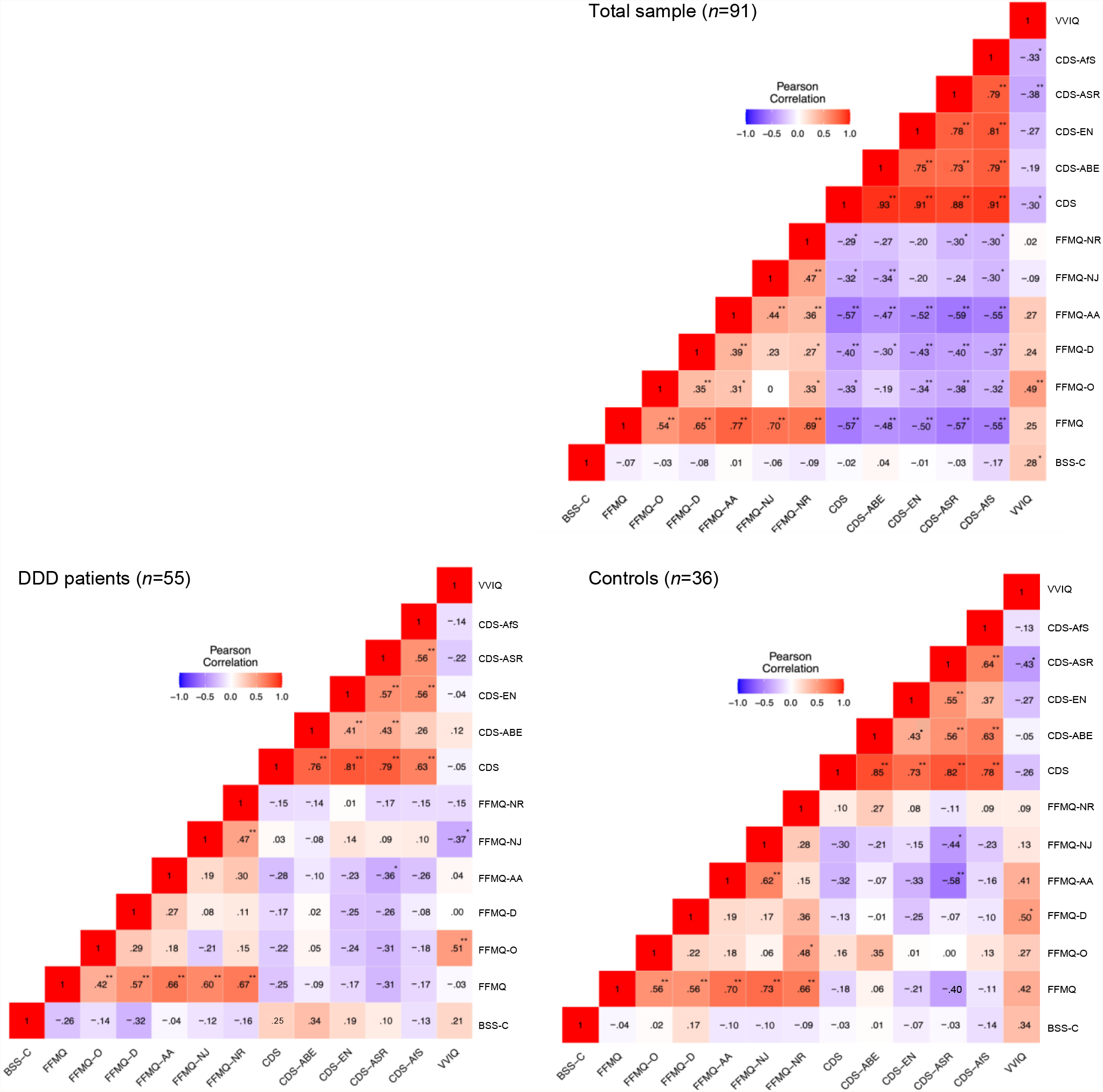
Correlations among all research variables. Data reported include Pearson correlation coefficients *Notes*. DDD = depersonalization-derealization disorder; BSS-C = *Brief Suggestibility Scale-Composite*; FFMQ-O = *Five Facet Mindfulness Questionnaire - Observing*; FFMQ-D = *Five Facet Mindfulness Questionnaire – Describing;* FFMQ-AA = *Five Facet Mindfulness Questionnaire – Acting with Awareness;* FFMQ-NJ = *Five Facet Mindfulness Questionnaire – Non-judging;* FFMQ-NR = *Five Facet Mindfulness Questionnaire – Non-reacting;* CDS-ABE = *Cambridge Depersonalization Scale* – Anomalous bodily experience; CDS-EN = *Cambridge Depersonalization Scale* – Emotional numbing; CDS-ASR = *Cambridge Depersonalization Scale* – Anomalous subjective recall; CDS-AfS = *Cambridge Depersonalization Scale* – Alienation from surroundings. \**p* < .01; \*\**p* < .001.

### 3.4 Responsiveness to suggestion and mindfulness

Our second moderation analysis tested the prediction that suggestibility would negatively relate to mindfulness (FFMQ-AA subscale) and that this effect would be more pronounced among patients. The overall model was non-significant, *F*(3, 87) = .63, *p* = .60, with weak non-significant effects for Group, *b* = -1.28, *t*(87) = -.71, *p* = .48, Acting with awareness, *b* = -.04, *t*(87) = -.62, *p* = .54, and their interaction, *b* = .03, *t*(87) = .37, *p* = .71. Correlation analyses between FFMQ-AA and BSS-C scores revealed a near-zero associations in the total sample, *r*(89) = .01, *p* = .90 [95% CI = -.19, .22], with similar effects in patients, *r*(53) = -.04, *p* = .76 [95% CI = -.30, .23], and controls, *r*(34) = -.10, *p* = .56 [95% CI = -.41, .24]. Similarly, correlations between total FFMQ and BSS-C scores, did not achieve significance in the total sample, *r*(89) = -.07, *p* = .50 [95% CI: -.27, .14], or controls, *r*(34) = -.04, *p* = .81 [95% CI: -.37, .29], although there was a trend toward a negative correlation in patients, *r*(53) = -.26, *p* = .056 [95% CI: -.49, .01]. The latter two effects did not significantly differ, *z* = -1.01, *p* = .31. Collectively, these results suggest that those DDD patients who were more suggestible were also less mindful, although this association did not differ from the corresponding effect in controls.

### 3.5 Exploratory analyses

Exploratory analyses investigated associations between the various research measures in the full sample and in patients and controls separately (**Figure 3**). Suggestibility and vividness of visual imagery (VVIQ) were significantly positively correlated in the total sample, *r*(89) = .28, *p* = .008 [95% CI = .08, .46], with a similar trend-level effect in controls, *r*(34) = .34, *p* = .043 [95% CI = .01, .60], but a weaker, non-significant effect in patients, *r*(53) = .21, *p* = .12 [95% CI = -.06, .45]. There was a trend-level effect in patients involving the CDS-ABE subscale, implying that those with more severe anomalous bodily experience scores were also more responsive to suggestions, *r*(53) = .34, *p* = .011 [95% CI = .08, .55]; this effect was near-zero and non-significant in the total sample, *r*(89) = .04, *p* = .71 [95% CI = -.17, .24], and in controls, *r*(34) = .00, *p* = .98 [95% CI = -.32, .33] (group correlation difference: *z* = 1.57, *p* = .12). A non-significant association between anxiety (GAD-7) and BSS-C scores was found in the total sample, *r*(89) = .08, *p* = .47 [95% CI: -.13, .28], and in controls alone, *r*(34) = -.06, *p* = .74 [95% CI: -.38, .28]. By contrast, in patients, there was trend-level positive correlation, *r*(53) = .29, *p* = .03 [95% CI: .03, .52], suggesting that those patients with more severe anxiety were also more responsive to suggestions, although these two group correlations did not significantly differ, *z* = -1.61, *p* = .11. Finally, exploratory analyses between suggestibility and mindfulness subscales revealed non-significant results in all cases. Beyond this, as seen **Figure 3**, the CDS and FFMQ, both total and subscales, are reliably negatively correlated in the total sample. This is most notable for the FFMQ-AA subscale with the CDS-ASR subscale, which is reliably negative in the total sample, as well as in patients and controls separately.

## 4 Discussion

On the basis of previous research highlighting elevated hypnotic suggestibility as a characteristic of dissociative psychopathology,^24,53^ this study investigated whether DDD is similarly characterized by aberrant responsiveness to direct verbal suggestions. The analyses revealed that DDD patients displayed non-significantly lower suggestibility than demographically matched controls. However, there were weak trends for responsiveness to suggestions to be associated with severity of depersonalization-derealization symptoms, particularly anomalous bodily experiences. In accordance with reports of attenuated mindfulness in high dissociation,^40,54^ patients with DDD also displayed lower mindfulness (FFMQ) than controls. These results indicate that DDD is not characterized by elevated direct verbal suggestibility and provide further insights into the aetiology and mechanisms of this condition and its status within the taxonomy of the dissociative disorders.

These results stand in stark contrast with the prediction that DDD patients, like those with other dissociative disorders, would be more responsive to direct verbal suggestions. However, the results do align with the possibility that elevated suggestibility is specifically linked to compartmentalization, and not detachment, symptoms and is not seen in anxiety disorders.^29^ Within the diagnosis of DDD, there is diverse symptomatology that overlaps with both anxiety and other dissociative disorders.^2,55-57^ In particular, most dissociative disorders such as dissociative amnesia and dissociative identity disorder are typified by compartmentalization symptoms including behavioural or emotional dysregulations, memory or identity disturbances, or functional neurological symptoms.^58^ By contrast, DDD is primarily characterised by detachment from one’s body, mental states, or sense of self (depersonalization) and/or from one’s surroundings (derealization).^57^ Recent work examining heterogeneity in DDD^13^ yielded evidence for five distinct classes of DDD patients: three comprising subtypes based on severity (Low severity, Moderate severity, High severity), and two subtypes differing on detachment and compartmentalization (High depersonalization, High dissociation) symptomatology.^3,4^ Accordingly, one interpretation of the present results is that elevated suggestibility is specific to a high dissociation (compartmentalization) subtype that possesses a more similar symptom profile to other dissociative disorders, or a high severity subtype, that also includes more severe anxiety, given the current trend towards more severe depersonalization-derealization symptoms as well as anxiety symptoms being associated with heightened suggestibility.

Another route for interpreting the apparent discrepancy between these results and evidence for elevated suggestibility in the dissociative disorders^24^ is the relationship between DDD and trauma. Whilst trauma is the primary antecedent of the dissociative disorders,^8-10^ precipitating factors for DDD are more varied and include substance use, depression, and panic^12-14^ with lower prevalence rates of self-reported childhood trauma.^12,15,59^ Accordingly, insofar as elevated direct verbal suggestibility is observed in dissociative, trauma and stressor-related disorders, such as PTSD^24-27,59,60^ and hypnotic suggestibility has been repeatedly shown to positively covary with posttraumatic symptoms^61-63^ elevated suggestibility is potentially specific to those suffering from trauma-related dissociative symptoms.^64^ At present, this interpretation is not discriminable from the view that elevated suggestibility is specific to compartmentalization symptomatology.

Previous research has demonstrated negative associations between mindfulness or metacognition and suggestibility^34,35,37,38^ implying that responsiveness to suggestion is supported by, or related to, aberrant metacognition pertaining to one’s intentions and the factors influencing their sense of agency.^65,66^ Similarly, preliminary research points to attenuated mindfulness in highly dissociative individuals^36,39,40,54^ and to attenuated intention awareness in germane populations.^67,68^ On the basis of this research, we examined whether suggestibility in DDD patients would be associated with, or moderated by, levels of mindfulness. In preliminary support of the former prediction, we observed a borderline significant negative correlation in patients, but not in controls or the total sample. This points to a potential role of lower mindfulness or poorer metacognition supporting greater responsiveness to suggestion in DDD patients that warrants greater attention in this population and in dissociative psychopathology more broadly.

The observation of marginally lower suggestibility in our sample of DDD patients is potentially attributable to our observation of attenuated mindfulness and imagery in this sample. Lower mindfulness in DDD patients, as observed here and suggested elsewhere,^54^ paired with elevated depersonalization-derealization symptoms, may be linked to reduced interoceptive awareness, an overall awareness and understanding of the body.^69^ It is possible that a certain level of awareness of the internal state of the body is necessary to experience suggested changes in behaviour and perception^70^ and a range of research points towards underactivity in brain areas associated with interoception in DDD.^71-74^ Similarly, this study replicates previous results^41^ suggesting that DDD patients display impairments in imagery compared to controls, particularly regarding self-related imagery. Responsiveness to suggestion does not reliably correlate with imagery, and the two seem to recruit distinct neurocognitive mechanisms.^75^ However, there is evidence that individuals with poor imagery are less responsive to suggestion, implying that some imagery capacity is necessary, but not sufficient, to respond to suggestions.^75,76^ We observed a significant positive correlation between suggestibility and vividness of visual imagery in the total sample, with a trend-level effect in controls but not in patients. This potentially aligns with previous research demonstrating evidence for a low dissociative, highly suggestible subtype with superior visual imagery.^77^ Taken together, these results suggest that aberrant interoceptive awareness and imagery in DDD may explain marginally lower suggestibility in this population.

These results have potential implications for therapeutic interventions in DDD. Insofar as suggestibility predicts treatment outcome with suggestion-based therapies (e.g., hypnotherapy^78,79^), the present results imply that these techniques are unlikely to be efficacious in this population. By contrast, given that we observed that DDD patients were characterized by reduced mindfulness, and mindfulness, particularly acting with awareness, tended to be negatively correlated with depersonalization-derealization symptoms, mindfulness-based treatments are probably a better target than suggestion-based treatments in DDD. Previous research has recommended training in mindfulness techniques as a potential therapeutic approach for DDD,^54^ with indications that mindfulness exercises, specifically mindful breathing, can immediately reduce present state depersonalization in patients with DDD (*d* = .65).^80^

Although these results should be interpreted with caution, they align with previous research showing that atypical suggestibility is specific to dissociative and germane disorders characterized by compartmentalization symptomatology,^4^ such as dissociative identity disorder^81^ and is positively related to functional and/or dissociative symptoms in functional neurological disorder.^32,82^ These results shed new light on the relationship between responsiveness to suggestion and dissociative psychopathology but should be considered in the context of multiple limitations. As the suggestibility assessment was online and unsupervised, we were unable to corroborate whether participants were complying with the experimental protocol, although use of this suggestibility scale has previously been shown to correlate with dissociative tendencies in a non-clinical sample.^11^ It is also possible that patients perceived the suggestibility assessment to index imagination and thus inferred that the procedure aimed to evaluate whether they were imagining their own symptoms.^4^ Accordingly, it may be valuable to measure suggestibility in DDD in a manner that doesn’t overtly reference imagination. Further, one notable confound of standardized suggestibility scales is that they include suggestions for dissociative and functional symptoms (i.e., amnesia, hallucinations, etc.) and it has been shown, for example, that FND patients are hyperresponsive to suggestions that modulate their symptoms.^24^ This suggests the possibility that elevated suggestibility in the dissociative disorders and FND is artefactual of the suggestion content of these scales. In turn, it will be imperative for future research on elevated suggestibility in dissociative psychopathology to include suggestions targeting non-dissociative, non-functional experience and symptoms (e.g., elevated positive affect). Conversely, it remains unexplored whether DDD patients would be more responsive to suggestions for the modulation of their detachment symptoms. If so, this may prove valuable in aiding the diagnosis of DDD as suggestive symptom induction is widely used to aid the diagnosis of FND.^83,84^ Another important consideration is the reason for particularly low prevalence rates of trauma in DDD specific samples. It is possible that this is due to a bias of referral pathways within clinical services: if patients report trauma, they will be referred to trauma focused services, leaving DDD specialist services and the research samples drawn from these predominantly seeing patients for whom these trauma referral pathways were not open. Lastly, studies exploring the links between dissociation and suggestibility often use the Dissociative Experiences Scale (DES).^7^ Since DDD may manifest as experiences of detachment and less so of compartmentalization, the CDS, as used in this study, is a valuable measure of this condition and the specific types of dissociation that DDD patients experience. However, future research on DDD and suggestibility should also include the DES to assess broader dissociative symptomatology and its relationship to suggestibility in DDD. Including this measure, along with the CDS, would also help to differentiate ostensible subtypes present within the DDD population^13^ and to evaluate our hypothesis that elevated suggestibility is specific to DDD patients experiencing compartmentalization symptoms. This and previous work^13^ suggests that DDD may not be best placed within the rubric of dissociative disorders and might be considered a distinct psychopathological syndrome.

## Data Availability

Data is fully available on OSF

https://osf.io/9pwzn/

## Acknowledgements

DBT was supported by Bial Foundation bursary 70/16.

